# Anemia and Its Associated Factors Among Women of Reproductive Age in Zambia: A Multilevel Mixed-Effects Analysis

**DOI:** 10.1101/2025.08.03.25332896

**Authors:** Given Moonga, Florence Mwila, James Muchinga, Mukuka Malasha, Christopher Kalumba, Esau Grecian Mbewe, Garikai Martin Membele, Kumbulani Mabanti

## Abstract

**Background:** Anemia remains a major global health crisis, affecting over 500 million women of reproductive age, with high burdens in resource-limited regions like Sub-Saharan Africa. Despite ongoing interventions such as iron supplementation programs, 49% of women of reproductive age in Zambia are anemic. Thus, the purpose of this study was to establish the national and subnational prevalence of anemia and identify its determinants among women of reproductive age in Zambia.

**Methods:** Data were drawn from the 2018 Zambia Demographic and Health Survey (ZDHS), a nationally representative survey employing a stratified two-stage cluster sampling design across 545 enumeration areas. A multilevel mixed-effects logistic regression model was used to identify individual-and community-level factors associated with anemia among women aged 15–49 years (n = 13,055). Four hierarchical models were constructed (null, individual-level, community-level, and full) to assess fixed and random effects, with model selection guided by AIC and BIC criteria. Spatial analysis was conducted using QGIS, incorporating displaced GPS coordinates in accordance with DHS protocols. All analyses applied sampling weights and assessed multicollinearity (VIFs < 5).

**Results:** The national prevalence of anemia among women of reproductive age was 31% (95% CI: 29– 33%), with the highest rates observed in Western (38%) and Lusaka (36%) provinces, and the lowest in Central Province (24%). In adjusted analyses, pregnancy (AOR = 1.76; 95% CI: 1.52– 2.03), HIV positivity (AOR = 2.21; 1.97–2.49), and breastfeeding (AOR = 1.15; 1.02–1.30) were significantly associated with increased odds of anemia. Conversely, being married (AOR = 0.78; 0.68–0.90) and age 25–29 years (AOR = 0.84; 0.71–0.97) were protective. Spatial mapping identified Western Province as a high-burden hotspot. Community-level variance was notable (ICC = 6%, MOR = 1.52), with 5% residual clustering persisting after adjusting for both individual and contextual factors, suggesting the influence of unmeasured ecological determinants.

**Conclusion:** Anemia remains a significant public health issue among Zambian women of reproductive age, shaped by both individual– and community-level factors. These findings highlight the need for integrated, targeted interventions focusing on high-risk groups in high-prevalence areas. Strengthening clinical services and implementing community-based strategies to address healthcare access and environmental determinants are essential to reducing the burden of anemia in Zambia.

## Introduction

Anemia is a condition characterized by a reduction in the number of red blood cells and the concentration of hemoglobin (Hb), which reduces the blood’s ability to carry adequate oxygen to meet the body’s physiological need [1]. The World Health Organization (WHO) defines anemia in non-pregnant women as a hemoglobin (Hb) concentration of less than 12 g/dL and in pregnanct women as less than 11 g/dL [1]. Anemia is a significant global public health concern, affecting over 500 million women of reproductive age (WRA) worldwide [2]. The World Health Organization (WHO) estimates that 37% of pregnant women and 30% of women aged 15-49 years worldwide are anemic [1].

The burden of anemia is disproportionately high in low-and middle-income countries (LMICs) and has negative impacts on maternal and child health which includes low birth weight, preterm birth, poor cognitive development perinatal and neonatal mortality, maternal morbidity and mortality and [3,4]. Additionally, anemia contributes to reduced productivity, cognitive impairment, and increased susceptibility to infections due to compromised immune function. It also affects nervous system, respiratory and circulatory system, skin mucous membrane, digestive system, endocrine system [5,6]. Africa and India bear the highest burden of anemia globally, with nearly half of all women affected. In these regions, anemia contributes to 40% of maternal deaths [7]. The greatest risk of anemia among women is seen in Sub-Saharan Africa (SSA), where 46% of pregnant women, and 39% of women of reproductive age are affected [8]. In Eastern Africa, anemia prevalence among women aged 15–49 years varies significantly, ranging from 19.2% in Rwanda to 49% in Zambia. The high prevalence of maternal anemia in Zambia, estimated at approximately 36.2%, underscores a significant public health concern in a country already facing challenges related to maternal mortality [9].

There are a number of factors attributed to anemia in pregnancy; these include micronutrient deficiencies of iron, folate, and vitamins A and B12 as well as infectious diseases such as HIV, tuberculosis, malaria and hookworm. Among these, iron deficiency contributes to around 50% of the anemia problem [4]. Iron deficiency is common among women of reproductive age largely because of their elevated demand for iron during pregnancy, lactation, menstrual blood loss, and nutritional deficiencies during their reproductive cycle [4,5]

Efforts to reduce anemia among women of reproductive age are being made in most countries including Zambia through iron supplementation and health education on nutrition. However, existing patterns of social inequalities in anemia over time among women of reproductive age are not well studied in most countries in the Sub-Saharan Region including Zambia. Existing disparities across places of residence and geographical areas are not well known [7,10]. For instance, a study in India revealed that anemia prevalence has increased significantly over a recent 7-year period, despite socioeconomic inequalities (by wealth, education and residence), having decreased over time [11].

The mixed-effects multivariable multilevel analysis becomes suitable to assess variation of anemia prevalence most studies use random effects to capture group specific variation in the study how they influence occurrence of anemia and these factors include community level and individual factors [12]. According to Teshale et al., (2020) anemia prevalence among rural women in Ethiopia was 25.1%, compared to 17.5% in urban areas and in his study, he found community factors like low household wealth was strongly associated with anemia in rural regions, suggesting that geographical and socioeconomic factors influence anemia prevalence.

Hence, this study aims to analyze the patterns of anemia prevalence among women of reproductive age in Zambia; and explore the determinants, and regional disparities. The study will contribute to informing national policy in the quest to meet the global commitment set by the World Health Organization (WHO) of achieving a 50% reduction of anemia in women of reproductive age by 2030.

## Methods

### Study design and setting

This study utilized a secondary data analysis approach based on the 2018 Zambia Demographic and Health Survey (ZDHS). The 2018 ZDHS was a nationally representative survey conducted by the Zambia Statistics Agency (ZamStats) in collaboration with the Ministry of Health (MOH). The cross-sectional survey and data collection spanned from 2018 to 2019. The main objective of the 2018 ZDHS was to collect up-to-date and comprehensive data on vital demographic and health indicators to inform national policy and planning. The ZDHS uses a stratified two-stage sampling design to ensure national, urban-rural, and provincial representativeness.

### Sampling and data measurement of ZDHS

In the first stage of the sampling design used in the 2018 ZDHS, enumeration areas (EAs) served as sampling clusters and were selected using probability proportional to size within each stratum. A total of 545 clusters were identified for inclusion. During the second stage, a household listing operation was conducted in these clusters, identifying an average of 133 households per cluster. Systematic sampling was then used to select 25 households from each cluster, culminating in a sample of 13,625 households. The detailed methodology for the ZDHS is well-documented in prior publications [14].

Anemia testing was done on all women of reproductive age (15–49 years old) from whom consent was obtained [14]. The missing data constituted only 4% and was, therefore, negligible since 96% of the selected women for the interviews responded.

### Explanatory variables (Risk Factors)

The study incorporated both individual-level and community-level variables available in the ZDHS dataset. Previous studies have attributed the prevalence of anemia to both individual and community-level factors

Individual-level factors encompassed demographic and health-related characteristics such as age, religion, marital status, educational attainment, wealth index, and gravidity. Community-level factors included variables such as residence type (urban or rural), geographic region, water source, and type of toilet facilities. These community-level variables describe populations living under similar environmental or infrastructural conditions. The classification of variables as either individual or community-level was based on the assumption of independence.

This study utilized secondary data that contained no personal identifiers. As such, the requirement for obtaining individual informed consent was waived by the University of Zambia Biomedical Research Ethics Committee (UNZABREC), which approved the study (Approval Number: 6300-2025).

### Statistical analysis

Statistical analysis was conducted using Stata 17 to generate descriptive statistics and address the research questions under investigation. Frequencies and percentages were used to summarize both individual– and community-level variables. Given that some regions with smaller populations were oversampled while others with larger populations were underrepresented, weighted frequencies and percentages were calculated to account for population size differences across regions. The detailed weighting methodology is outlined in the ZDHS 2018 report [14].

To assess the factors influencing anemia among women, a bivariate logistic regression was performed to identify significant variables for inclusion in the multivariate standard logistic regression and mixed-effects multivariable multilevel logistic regression models. Age, marital status, breastfeeding status, pregnancy status, HIV test results, religion, province, sex of the household head, gravidity, and the number of children ever born in the preceding 5 years were statistically significantly associated with anemia. Missing observations in the anemia and HIV variables were omitted, and inconclusive responses in the HIV variable were also omitted. This resulted in a reduction of observations from 13,625 to 13,055. Gravidity was dropped due to its high correlation with marital status and age.

To select between the multivariate standard logistic regression and the mixed-effects multivariable multilevel logistic regression model, a log-likelihood ratio test was conducted. A model with a higher log-likelihood value is preferred [15].

### Standard logistic model

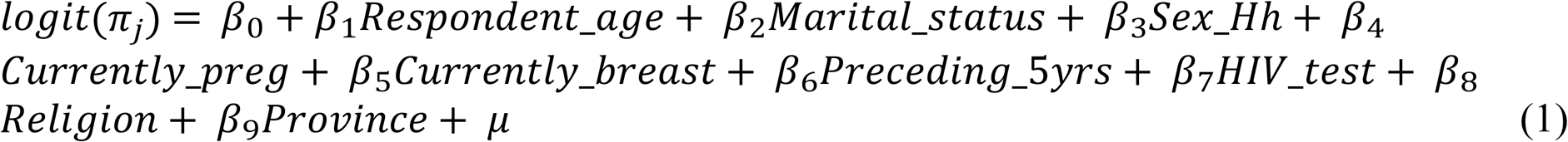

### Mixed-effects multilevel model

A mixed effects multilevel model is a standard logistic regression model, amended to have random effects for each EA. Defining π_*ij*_ = Pr (*Anemia*_*status*_ = 1), we have

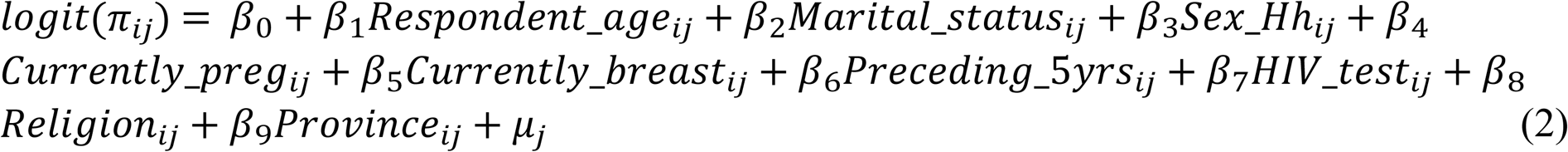

for *j* = 1, …, 545 EAs, with *i* = 1, …, *n*_*j*_ women in EA *j*.

### Model selection

A log-likelihood ratio test was conducted to select between the standard logistic regression and the mixed-effects multilevel logistic regression model. As a rule of thumb can be seen that a model with a higher log-likelihood value is preferred [15].

### Mixed-effects logistic regressions

The fit of the models are compared by relying on Akaike Information Criterion (AIC) and the Bayesian Information Criterion (BIC) and are summarised in Table 2. As can be seen that the model with the lowest value describes the data best [16].

**Table 1:** Standard logistic and Mixed effects multilevel regressions.

| Model | Loglikelihood Ratio |
| --- | --- |
| Standard logistic regression | -7811 |
| **Mixed effects multilevel regression** | **-7757** |

**Table 2:** Estimation results: AIC and BIC.

| Model | AIC | BIC |
| --- | --- | --- |
| Model 1 | 15868 | 15868 |
| Model 2 | 15586 | 15683 |
| Model 3 | 15838 | 15942 |
| **Model 4** | **15561** | **15755** |

We specified 4 distinct models, aiming to identify the risk factors at different levels, model 1 included no predictors to assess community-level variation in anemia prevalence; model 2 added individual-level factors; model 3 included community-level factors; and Model 4 combined both individual– and community-level factors. In more detail, the differences between the models are summarized in Table 3.

**Table 3.** Specification of estimated models.

| Model term | Model 1 | Model 2 | Model 3 | Model 4 |
| --- | --- | --- | --- | --- |
| $\beta_0$ | y<sup>a</sup> | y | y | y |
| $\beta_1$*Respondent_age* | n<sup>b</sup> | y | n | y |
| $\beta_2$*Marital_status* | n | y | n | y |
| $\beta_3$*Sex_Hh* | n | y | n | y |
| $\beta_4$*Currently_preg* | n | y | n | y |
| $\beta_5$*Currently_breast* | n | y | n | y |
| $\beta_6$*Preceding_5yrs* | n | y | n | y |
| $\beta_7$*HIV_test* | n | y | n | y |
| $\beta_8$*Religion* | n | y | n | y |
| $\beta_9$*Province* | n | n | y | y |

<sup>a</sup> Yes (y) and <sup>b</sup>No (n)

The fixed-effects results were expressed as odds ratios (ORs) with 95% confidence intervals (CIs), while adjusted odds ratios (AORs) with 95% CIs were calculated to identify independent factors associated with anemia. Statistical significance was determined using a p-value threshold of <0.05. To assess multicollinearity, variance inflation factors (VIFs) were computed, with variables considered to have low collinearity if their VIF values were below [17]

Random effects, capturing variations between clusters, were assessed using the Intra-Cluster Correlation Coefficient (ICC), Percentage Change in Variance (PCV), and Median Odds Ratio (MOR). The ICC quantified the proportion of total variability attributable to clustering effects, while the MOR converted unexplained cluster-level variance into the OR scale, facilitating the interpretation of heterogeneity. The PCV measured the proportion of total variance explained by individual– and community-level factors. These measures collectively informed the suitability of the model specification. These measures were computed using the following formulas:

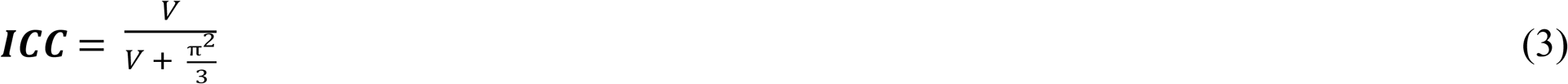

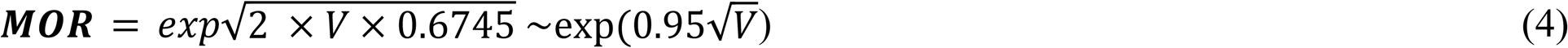

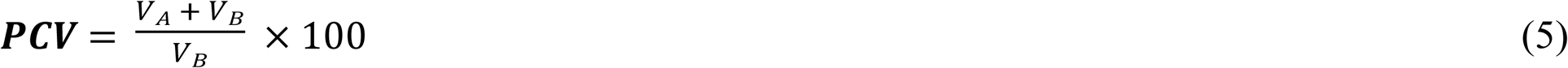

Where V represents the estimated variance of clusters, *V*_*A*_ is the variance in the initial model, and *V*_*B*_ is the variance in subsequent models with additional terms. The multi-level analysis was well suited for our study given the correlated nature of our data. It evaluates how variables at different levels influence the dependent variable while addressing biases introduced by clustering [18]. By producing corrected standard errors, confidence intervals, and significance tests, multilevel models yield reliable parameter estimates.

The results of the fourth model, selected based on the AIC and BIC, indicated that age group, marital status, sex of the household head, pregnancy status, breastfeeding status, number of children born in the preceding five years, HIV test results, residence, and religion were significantly associated with anemia among women.

### Spatial mapping

The ZDHS data is georeferenced, enabling spatial analysis to be carried out. To maintain the confidentiality of respondents, ZDHS has a georeferenced data release policy, which provides two levels of protection. First, data from the same enumeration area (EA) are aggregated to a single-point coordinate. Second, a GPS coordinate displacement process is applied, which masks the true locations. Urban clusters were displaced a distance of up to 2 km (0-2 km), rural clusters up to 5 km (0-5 km), and an additional randomly selected 1% of rural clusters are displaced up to ten kilometers (0-10 km) [14].

A spatial analysis was conducted to assess the prevalence of anemia among women of reproductive age (WRA) in Zambia (2018) using geospatial data from the Zambia Demographic and Health Survey (ZDHS). The shapefile for Zambia’s administrative boundaries containing the prevalence of anemia in percentages was obtained from the ZDHS website. The GIS data was imported into QGIS for spatial visualization. Anemia prevalence was categorized into seven classes using a colour gradient, with yellow indicating the lowest prevalence and red the highest prevalence. Labels for each province were added for better interpretation. The prevalence was mapped across Zambia’s province, highlighting spatial disparities in anemia levels among WRA.

## Results

### Sociodemographic characteristic

Table 1 presents the descriptive statistics of the study participants. The analysis included data on a total of 13,055 women from 545 clusters across 10 provinces. The median age (IQR) of the participants was 27 (20 to 36) years. Over half (56%) of the women were married, and 8% had no formal education. About half (54%) of the women in the study were rural residents, and 71% identified as protestant Christians. About 8% of the women were pregnant at the time of the survey, while 23% were breastfeeding and 14% were HIV positive. Furthermore, nearly three-quarters (73%) of the households were male-headed, 72% had access to improved water sources while 57% had unimproved toilet facilities.

Table 4 shows the variation of anemia across different Sociodemographic and health-related characteristics among study participants. The estimated anemia prevalence among women of reproductive age was 31%. Significant regional variations were observed, Lusaka and Western had the highest prevalence (36% and 38%, respectively), while Central province had the lowest (24%). Younger women, particularly those aged 15-19, had the highest anemia prevalence (33%), while older women (40-49 years) also had elevated prevalence (33%). Single women exhibited higher anemia prevalence (34%) compared to married women (28%). The prevalence of anemia among pregnant women was higher (41%) compared to the women who were not pregnant (30%).

**Table 4.** Descriptive statistics and anemia prevalence.

| Covariates | Weighted Frequency | Weighted percent | Anemic | Non-Anemic | Weighted Frequency | *P-value |
| --- | --- | --- | --- | --- | --- | --- |
| **Age Groups** |  |  |  |  |  |  |
| 15-19 | 2878 | 22% | 958 | 1920 | 33% (31 to 36) | 0.005 |
| 20-24 | 2626 | 20% | 741 | 1885 | 28% (26 to 31) |  |
| 25-29 | 2120 | 16% | 611 | 1509 | 29% (26 to 32) |  |
| 30-39 | 3380 | 26% | 1058 | 2321 | 31% (29 to 34) |  |
| 40-49 | 2051 | 16% | 679 | 1373 | 33% (31 to 36) |  |
| **Marital Status** |  |  |  |  |  |  |
| Single | 4061 | 31% | 1377 | 2684 | 34% (32 to 36) | <0.001 |
| Married | 7281 | 56% | 2060 | 5220 | 28% (27 to 30) |  |
| Divorced | 1713 | 13% | 610 | 1104 | 36% (33 to 38) |  |
| **Education Level** |  |  |  |  |  |  |
| Higher education | 698 | 5% | 246 | 451 | 35% (31 to 40) | 0.323 |
| Secondary | 5532 | 42% | 1721 | 3811 | 31% (29 to 34) |  |
| Primary | 5831 | 45% | 1773 | 4059 | 30% (29 to 32) |  |
| No education | 994 | 8% | 307 | 687 | 31% (27 to 35) |  |
| **Sex of household head** |  |  |  |  |  |  |
| Male | 9514 | 73% | 2840 | 6674 | 30% (28 to 31) | <0.001 |
| Female | 3541 | 27% | 1207 | 2334 | 34% (32 to 36) |  |
| **Residence** |  |  |  |  |  |  |
| Urban | 6013 | 46% | 1931 | 4082 | 32% (30 to 35) | 0.163 |
| Rural | 7042 | 54% | 2116 | 4926 | 30% (29 to 32) |  |
| **Region** |  |  |  |  |  |  |
| Central | 1133 | 9% | 269 | 864 | 24% (21 to 27) | <0.001 |
| Copperbelt | 2042 | 16% | 600 | 1442 | 29% (26 to 33) |  |
| Eastern | 1541 | 12% | 425 | 1116 | 28% (24 to 31) |  |
| Luapula | 967 | 7% | 286 | 681 | 30% (27 to 33) |  |
| Lusaka | 2658 | 20% | 945 | 1713 | 36% (32 to 39) |  |
| Muchinga | 742 | 6% | 205 | 537 | 28% (22 to 33) |  |
| Northern | 1039 | 8% | 289 | 749 | 28% (24 to 32) |  |
| North western | 690 | 5% | 224 | 467 | 32% (28 to 37) |  |
| Southern | 1486 | 11% | 520 | 966 | 35% (31 to 40) |  |
| Western | 757 | 6% | 284 | 473 | 38% (34 to 41) |  |
| **Religion** |  |  |  |  |  |  |
| Catholic | 2245 | 17% | 682 | 1563 | 30% (28 to 33) | <0.001 |
| Protestant | 10587 | 81% | 3273 | 7314 | 31% (29 to 32) |  |
| Muslim | 61 | 1% | 13 | 48 | 21% (13 to 33) |  |
| Other | 162 | 1% | 79 | 83 | 49% (39 to 58) |  |
| **Wealth Index** |  |  |  |  |  |  |
| Poorest | 2334 | 18% | 702 | 1632 | 30% (28 to 32) | 0.548 |
| Poorer | 2307 | 18% | 704 | 1602 | 31% (28 to 33) |  |
| Middle | 2383 | 18% | 725 | 1658 | 30% (28 to 33) |  |
| Richer | 2906 | 22% | 894 | 2013 | 31% (28 to 34) |  |
| Richest | 3125 | 24% | 1022 | 2103 | 33% (30 to 36) |  |
| **Currently Pregnant** |  |  |  |  |  |  |
| Yes | 1074 | 8% | 442 | 632 | 41% (36 to 46) | <0.001 |
| No | 11981 | 92% | 3605 | 8376 | 30% (29 to 31) |  |
| **Currently Breastfeeding** |  |  |  |  |  |  |
| Yes | 2985 | 23% | 822 | 2163 | 28% (26 to 30) | <0.001 |
| No | 10070 | 77% | 3225 | 6845 | 32% (30 to 34) |  |
| **Gravidity of women (Children ever born)** |  |  |  |  |  |  |
| 0 | 3191 | 24% | 1106 | 2085 | 35% (32 to 37) | <0.001 |
| 1-3 | 5472 | 42% | 1657 | 3815 | 30% (28 to 32) |  |
| 4+ | 4392 | 34% | 1284 | 3108 | 29% (28 to 31) |  |
| **Children ever born in the preceding 5 years** |  |  |  |  |  |  |
| 0 | 6013 | 46% | 2053 | 3960 | 29% (26 to 31) | <0.001 |
| 1 | 4820 | 37% | 1381 | 3439 | 28% (25 to 30) |  |
| 2+ | 2222 | 17% | 613 | 1609 | 35% (32 to 37) |  |
| **Smoking** |  |  |  |  |  |  |
| Yes | 346 | 3% | 89 | 257 | 26% (21 to 31) | 0.059 |
| No | 12709 | 97% | 3958 | 8751 | 31% (30 to 33) |  |
| **HIV test** |  |  |  |  |  |  |
| Positive | 1863 | 14% | 843 | 1020 | 45% (42 to 49) | <0.001 |
| Negative | 11192 | 85% | 3204 | 7988 | 29% (27 to 30) |  |
| **Getting medical help for self:**<br>**distance to health facility** |  |  |  |  |  |  |
| Big problem | 3794 | 29% | 1189 | 2606 | 31% (30 to 33) | 0.715 |
| Not a big problem | 9261 | 71% | 2858 | 6402 | 31% (29 to 33) |  |
| **Have mosquito bed net for sleeping** |  |  |  |  |  |  |
| Yes | 10601 | 81% | 3236 | 7365 | 33% (30 to 36) | 0.082 |
| No | 2454 | 19% | 811 | 1643 | 31% (30 to 33) |  |
| **Type of water source** |  |  |  |  |  |  |
| Improved | 9425 | 72% | 2954 | 6571 | 31% (30 to 33) | 0.503 |
| Unimproved | 3291 | 25% | 984 | 2307 | 30% (28 to 32) |  |
| Other | 339 | 3% | 109 | 230 | 32% (26 to 39) |  |
| **Type of toilet** |  |  |  |  |  |  |
| Improved | 4211 | 32% | 1317 | 2895 | 31% (29 to 34) | 0.847 |
| Unimproved | 7425 | 57% | 2275 | 5149 | 31% (29 to 32) |  |
| Open | 1095 | 8% | 353 | 742 | 32% (29 to 36) |  |
| Others | 324 | 3% | 102 | 222 | 31% (25 to 38) |  |

\*p-values generated from Pearson chi2-test

Factors such as marital status, HIV positivity, currently pregnant, currently breastfeeding, and gravidity were highly correlated (*p* = 0.001) with anemia prevalence while access to medical help, residence and sanitation showed no significant association.

Anemia prevalence was categorized into seven groups, with yellow indicating lower prevalence and red representing higher prevalence. The map highlights spatial disparities in anemia prevalence across Zambia, with Central province having low anemia prevalence and Western province having the highest prevalence in the country as shown in Fig 1.

**Fig. 1.**
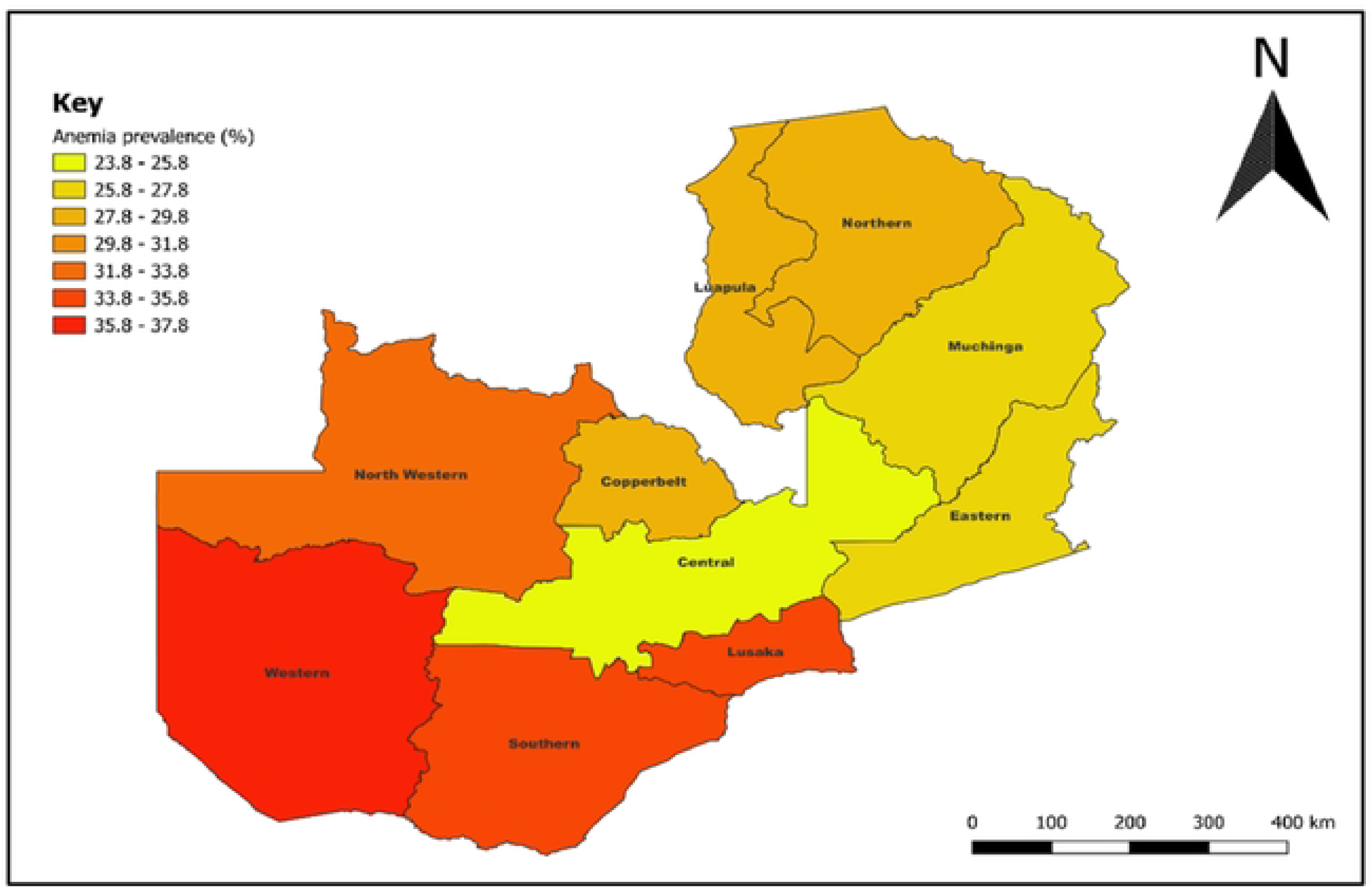
Map of Zambia showing the prevalence of anemia at the subnational level. Spatial Data Repository, the Demographic and Health Surveys Program. ICF International. Funded by **the** United States Agency for International Development (USAID). Available from spatialdata.dhsprogram.com. [Accessed 16 January 2025)

### Fixed effect

Table 5 shows the results of the fixed effect model. Women aged 25–29 years old were 16% less likely to be anemic compared with women in the youngest age group (15–19 years old) (AOR=0.84; 95% CI 0.71 to 0.97). The married women were 0.78 times less likely to be anemic than women who never married (AOR= 0.78; 95 % CI 0.68 to 0.90). The odds of anemia were higher in women who were pregnant (AOR=1.76; 95 % CI 1.52 to 2.03) compared with those who were not pregnant. Women who were currently breastfeeding were 15% (AOR=1.15; 95% CI 1.02 to 1.30) more likely to be anemic. Women who gave birth to one child in the preceding 5 years of the survey were at low risk of having anemia (AOR= 0.84; 95% CI 0.76 to 0.94). In this study, women who were HIV positive had a twofold increased odds of having anemia compared with women classified as HIV negative (AOR=2.21; 95% CI (1.97 to 2.49). Higher odds of anemia were observed in Copperbelt(AOR =1.33; 95% CI 1.06 to 1.68), Eastern (AOR=1.27; 95% CI 1.01 to 1.61), Lusaka (AOR=1.3; 95% CI 1.03 to 1.66), Luapula (AOR = 1.52; 95% CI 1.21 to 1.90), Northern (AOR =1.31; 95% CI 1.03 to 1.67), Northwestern (AOR =1.74; 95% CI 1.36 to 2.22), Southern(AOR =1.52; 95% CI 1.20 to 1.91) and Western provinces (AOR =1.99; 95% CI 1.56 to 2.54) compared with Central province.

**Table 5.** Multilevel mixed effects logistic regression analysis for determinant factors associated with anemia among women of reproductive age.

| **Variables** | **Model 1** | **Model 2**<br>**Individual-level**<br>**factors OR (95% CI)** | **Model 3**<br>**Community-level**<br>**factors OR (95% CI)** | **Model 4**<br>**Individual + community-level**<br>**factors**<br>**OR (95% CI)** |
| --- | --- | --- | --- | --- |
| **Age Groups** |  |  |  |  |
| 15-19 |  | 1 |  | 1 |
| 20-24 |  | 0.89 (0.78 to 1.02) * |  | 0.88 (0.77 to 1.01) * |
| 25-29 |  | 0.84 (0.72 to 0.99) ** |  | 0.83 (0.71 to 0.97) ** |
| 30-39 |  | 0.97 (0.84 to 1.13) |  | 0.95 (0.82 to 1.11) |
| 40-49 |  | 1.04 (0.88 to 1.23) |  | 1.02 (0.86 to 1.20) |
| **Marital Status** |  |  |  |  |
| Single |  | 1 |  | 1 |
| Married |  | 0.76 (0.66 to 0.87) *** |  | 0.78 (0.68 to 0.90) *** |
| Divorced |  | 0.89 (0.76 to 1.04) |  | 0.91 (0.77 to 1.07) |
| **Sex of household head** |  |  |  |  |
| Male |  | 1 |  | 1 |
| Female |  | 1.04 (0.94 to 1.16) |  | 1.03 (0.93 to 1.14) |
| **Currently Pregnant** |  |  |  |  |
| No |  | 1 |  | 1 |
| Yes |  | 1.77 (1.53 to 2.04) *** |  | 1.76 (1.52 to 2.03) *** |
| **Currently Breastfeeding** |  |  |  |  |
| No |  | 1 |  | 1 |
| Yes |  | 1.16 (1.03 to 1.31) ** |  | 1.15 (1.02 to 1.30) ** |
| **Children ever born in the preceding 5 years** |  |  |  |  |
| 0 |  | 1 |  | 1 |
| 1 |  | 0.85 (0.76 to 0.95) *** |  | 0.84 (0.76 to 0.94) *** |
| 2+ |  | 0.91 (0.78 to 1.06) |  | 0.991 (0.78 to 1.06) |
| **HIV test** |  |  |  |  |
| Negative |  | 1 |  | 1 |
| Positive |  | 2.22 (1.98 to 2.50) *** |  | 2.21 (1.97 to 2.49) *** |
| **Religion** |  |  |  |  |
| Catholic |  | 1 |  | 1 |
| Protestant |  | 1.02 (0.92 to 1.14) |  | 1.01 (0.90 to 1.12) |
| Other |  | 1.30 (0.93 to 1.82) |  | 1.27 (0.91 to 1.78) |
| **Region** |  |  |  |  |
| Central |  |  | 1 | 1 |
| Copperbelt |  |  | 1.35 (1.08 to 1.69) *** | 1.33 (1.06 to 1.68) ** |
| Eastern |  |  | 1.20 (0.96 to 1.50) | 1.27 (1.01 to 1.61) ** |
| Luapula |  |  | 1.25 (0.99 to 1.57) * | 1.31 (1.03 to 1.66) ** |
| Lusaka |  |  | 1.52 (1.22 to 1.89) *** | 1.52 (1.21 to 1.90) *** |
| Muchinga |  |  | 1.05 (0.82 to 1.34) | 1.15 (0.89 to 1.47) |
| Northern |  |  | 1.20 (0.95 to 1.51) | 1.31 (1.03 to 1.67) ** |
| North western |  |  | 1.62 (1.28 to 2.06) *** | 1.74 (1.36 to 2.22) *** |
| Southern |  |  | 1.48 (1.18 to 1.86) *** | 1.52 (1.20 to 1.91) *** |
| Western |  |  | 1.99 (1.57 to 2.52) *** | 1.99 (1.56 to 2.54) *** |
| **Random effects** |  |  |  |  |
| Community level variance (SE) | 0.19<br>(0.03) | 0.20 (0.03) | 0.15 (0.02) | 0.16 (0.02) |
| P value | <0.000 | <0.000 | <0.000 | <0.000 |
| DIC (−2log likelihood) | -7931.79 | -7778.51 | -7905.97 | -7756.62 |
| ICC (%) | 5.55 | 5.83 | 4.50 | 4.90 |
| Explained variation – PCV (%) | Reference | -5.17 | 24.78 | 13.92 |
| MOR | 1.52 | 1.54 | 1.45 | 1.48 |

Note: \*\*\* p < 0.01, \*\* p < 0.05, \* p < 0.1. DIC, Deviance Information Criterion; ICC, Intra-Cluster Correlation Coefficient; MOR, Median OR; PCV, Percentage Change in Variance.

### Random effects

The results of the random effects model are shown in Table 5 showed that the prrevalence rate of anemia varied across communities (p < 0.001). In other words, the anemia prevalence rate was not similarly distributed across the communities. About 6% of the variance in the odds of anemia in women could be attributed to community-level factors, as calculated by the ICC based on estimated intercept component variance. After adjusting for individual-level and community-level factors, the variation in anemia across communities remained statistically significant. About 5% of the odds of anemia variation across communities was observed in the full model (model 4). In the null model, the MOR for anemia was 1.52, indicating that there was a variance between communities (clustering) (1.52 times larger than the reference (MOR = 1). When both individual and community factors were included in the model, the unexplained community variation in anemia decreased to MOR of 1.48. This indicates that in the full model the effects of clustering are still statistically significant when we considered both individual and community factors.

## Discussion

Approximately 31% of women of reproductive age (WRA) were anaemic in the current study, and in comparison with WHO thresholds, it confirms that anemia remains a moderate public health concern in Zambia. This study investigated the prevalence, spatial patterns, and determinants of anemia among WRA in Zambia. The prevalence of anaemia in Zambia is slightly above the global average of 29.9% [19]. The prevalence reported in this study is also higher than those observed in Ethiopia (23%) and Rwanda (19.2%) [20], indicating comparatively greater vulnerabilities among Zambian women, potentially driven by biological factors such as frequent blood loss and gestational demands, as well as socio-environmental factors such as food insecurity and limited access to healthcare [20]. Nevertheless, Zambia’s anemia prevalence remains lower than that recorded in countries such as the Democratic Republic of the Congo (42.4%), Burundi (38.5%), South Sudan (35.6%), the Central African Republic (46.8%), Tanzania (38.9%), Uganda (32.8%), Congo-Brazzaville (48.8%), and Angola (44.5%) [19].

Spatial analysis revealed significant geographic variation in anemia prevalence, which may reflect provincial-level differences in dietary patterns, communicable disease burden, and healthcare access. Additionally, environmental determinants such as access to clean water and sanitation were also noted to influence anemia risk through increased exposure to soil-transmitted helminths [21]. Western province recorded the highest anemia prevalence (38%), followed closely by Lusaka province (36%). The elevated burden in Western province may stem from widespread poverty and reliance on subsistence farming, which limits access to iron-rich and diverse diets. In Lusaka, a study attributed anemia risk to poor vegetable intake and low socioeconomic status, particularly in informal urban settlements [22]. Consistent with previous findings in low– and middle-income countries (LMICs), this study also found that women residing in rural areas were more likely to be anemic than their urban counterparts. Contributing factors may include limited access to nutritious foods, lower socioeconomic conditions, and poor sanitation, all of which elevate disease exposure and contribute to increased anemia risk [23].

The multilevel mixed effects model indicated that both individual– and community-level factors accounted for approximately 14% of the variation in anemia prevalence. Age was a significant predictor; women aged 25–29 years had lower odds of anemia than those aged 15–19 years, consistent with findings from Ethiopia and Benin [21]. This could reflect lower fertility rates and better health-seeking behaviors among women in their mid-20s, including higher adherence to antenatal supplementation [5]. However, contradictory findings from Burkina Faso suggest an increased risk of severe anemia in this age group, potentially due to ongoing physiological development and menstrual iron loss compounded by inadequate dietary intake [24]. Marital status was also found to be a protective factor against anemia. Married women were less likely to be anemic than never-married women, likely due to increased access to financial, emotional, and nutritional support within marital households [25]. Such support may improve dietary adequacy and access to healthcare services, indirectly reducing anemia risk.

Pregnancy was positively associated with anemia, echoing evidence from Sub-Saharan Africa and LMICs, including Ethiopia, Mali, and Gambia [26]. The physiological changes that occur during pregnancy including blood volume expansion, fetal growth, and placental development elevate iron requirements, which may not be met through diet alone, particularly in resource-constrained settings [25]. Nonetheless, studies from Rwanda have reported no significant association between pregnancy and anemia, underscoring the importance of country-specific contextual factors [27]. Breastfeeding was similarly associated with a higher risk of anemia, consistent with studies from Ethiopia, India, and the Democratic Republic of Congo [19]. The increased nutrient demands during lactation, particularly for vitamins such as thiamine, riboflavin, B6, B12, A, and iodine, may not be adequately met among Zambian women, especially in rural areas where dietary diversity is limited [19]. These deficiencies can compromise hemoglobin synthesis and maternal iron stores during the postpartum period.

Women who had given birth to one child in the past five years were less likely to be anemic than those who had not given birth during that period. This finding aligns with evidence from Zambia and East Africa [28] and may reflect increased engagement with maternal health services and heightened health awareness during and after pregnancy. However, it contrasts with evidence from Mali and Ethiopia, where high gravidity and frequent births were associated with elevated anemia risk due to repeated blood loss, nutritional depletion, and parasitic infection [29]. These contrasting results suggest that the relationship between parity and anemia is complex and context-dependent. Lastly, HIV-positive status was positively associated with anemia, a finding consistent with studies from Ethiopia [21]. The elevated risk may be explained by HIV-related bone marrow suppression, chronic inflammation, and increased susceptibility to opportunistic infections that interfere with red blood cell production and iron utilization.

## Conclusion

Anemia among women of reproductive age in Zambia persists as a significant public health concern, driven by a combination of biological, socio-demographic, and community-level factors. This study underscores the importance of multilevel and spatial approaches in revealing hidden geographic and structural disparities. Effective responses will require integrated interventions that move beyond individual supplementation to address broader systemic issues such as food insecurity, access to care, and environmental health risks. Future research should explore these unmeasured contextual drivers and evaluate the impact of locally tailored, multisectoral strategies.

## Data Availability

Data Repository, the Demographic and Health Surveys Program

NA

## APPENDIX

**Table 5.** Bivariate and multivariable logistic models.

| Variable | Crude Odds (95% CI) | p value | Adjusted Odds (95% CI) | p value |
| --- | --- | --- | --- | --- |
| **Age Groups** |  |  |  |  |
| 15-19 | 1 |  | 1 |  |
| 20-24 | 0.84 (0.75-0.95) | 0.003 | 0.89 (0.78-1.01) | 0.074 |
| 25-29 | 0.80 (0.71-0.90) | 0.001 | 0.86 (0.74-0.10) | 0.047 |
| 30-39 | 0.93 (0.84-1.03) | 0.184 | 0.97 (0.84-1.13) | 0.715 |
| 40-49 | 1.01 (0.89-1.13) | 0.887 | 1.03 (0.88-1.21) | 0.722 |
| **Marital Status** |  |  |  |  |
| Single | 1 |  | 1 |  |
| Married | 0.81 (0.74-0.88) | <0.001 | 0.78 (0.68-0.89) | <0.001 |
| Divorced | 1.10 (0.98-1.24) | 0.177 | 0.90 (0.77-1.05) | 0.193 |
| **Education Level** |  |  |  |  |
| No or primary education | 1 |  | NA |  |
| Higher education | 0.990506 (0.92-1.07) | 0.863 |  |  |
| **Sex of household head** |  |  |  |  |
| Male | 1 |  | 1 |  |
| Female | 1.24 (1.14-1.35) | <0.001 | 1.04 (0.94-1.45) | 0.488 |
| **Residence** |  |  |  |  |
| Urban | 1 |  | 1 |  |
| Rural | 0.98 (0.91-1.06) | 0.638 | 1.22 (1.03-1.22) | 0.009 |
| **Region** |  |  |  |  |
| Central | 1 |  | 1 |  |
| Copperbelt | 1.36 (1.15-1.16) | <0.001 | 1.39 (1.17-1.64) | <0.001 |
| Eastern | 1.19 (1.00-1.41) | 0.040 | 1.25 (1.05-1.48) | 0.010 |
| Luapula | 1.25 (1.05-1.49) | 0.011 | 1.30 (1.09-1.55) | 0.003 |
| Lusaka | 1.55 (1.32-1.81) | <0.001 | 1.60 (1.35-1.88) | <0.001 |
| Muchinga | 1.08 (0.90-1.29) | 0.416 | 1.16 (0.97-1.39) | 0.110 |
| Northern | 1.20 (1.01-1.43) | 0.042 | 1.30 (1.09-1.56) | 0.004 |
| North western | 1.63 (1.37-1.95) | <0.001 | 1.73 (1.45-2.08) | <0.001 |
| Southern | 1.47 (1.24-1.75) | <0.001 | 1.50 (1.26-1.78) | <0.001 |
| Western | 1.99 (1.67-2.37) | <0.001 | 1.97 (1.64-2.35) | <0.001 |
| **Religion** |  |  |  |  |
| Catholic | 1 |  | 1 |  |
| Protestant | 1.03 (0.93-1.14) | 0.501 | 1.00 (0.90-1.11) | 0.998 |
| Other | 1.38 (1.01-1.89) | 0.043 | 1.30 (0.94-1.79) | 0.112 |
| **Wealth Index** |  |  |  |  |
| Poorest | 1 |  | NA |  |
| Poorer | 1.00 (0.89-1.12) | 0.954 |  |  |
| Middle | 0.95 (0.84-1.06) | 0.337 |  |  |
| Richer | 0.94 (0.83-1.05) | 0.279 |  |  |
| Richest | 1.00 (0.90-1.13) | 0.938 |  |  |
| **Currently Pregnant** |  |  |  |  |
| No | 1 |  | 1 |  |
| Yes | 1.46 (1.28-1.66) | <0.001 | 1.73 (1.50-1.99) | <0.001 |
| **Currently Breastfeeding** |  |  |  |  |
| No | 1 |  | 1 |  |
| Yes | 0.88 (0.81-0.97) | <0.001 | 1.14 (1.01-1.29) | <0.001 |
| **Gravidity of women (Children ever born)** |  |  |  |  |
| 0 | 1 |  | NA |  |
| 1 to 3 | 0.84 (0.75-0.91) | <0.001 |  |  |
| 4+ | 0.81 (0.74-0.90) | <0.001 |  |  |
| **Children ever born in the preceding 5 years.** |  |  |  |  |
| 0 | 1 |  | 1 |  |
| 1 | 0.83 (0.72-0.86) | <0.001 | 0.84 (0.76-0.94) | 0.001 |
| 2+ | 0.82 (0.71-0.89) | <0.001 | 0.91 (0.78-1.06) | 0.223 |
| **Smoking** |  |  |  |  |
| No | 1 |  | NA |  |
| Yes | 0.89 (0.71-1.11) | 0.310 |  |  |
| **HIV test** |  |  |  |  |
| Negative | 1 |  | 1 |  |
| Positive | 2.05 (1.85-2.27) | <0.001 | 2.14 (1.92--2.49) | <0.001 |
| **Getting medical help for self: distance to health facility** |  |  |  |  |
| Big problem | 1 |  | NA |  |
| Not a big problem | 0.96 (0.89-1.04) | 0.315 |  |  |
| **Have mosquito bed net for sleeping** |  |  |  |  |
| No | 1 |  | NA |  |
| Yes | 0.97 (0.88-1.07) | 0.607 |  |  |
| **Type of water source** |  |  |  |  |
| Improved | 1 |  | NA |  |
| Unimproved | 1.01 (0.93-1.10) | 0.835 |  |  |
| Other | 1.08 (0.85-1.36) | 0.534 |  |  |
| **Type of toilet** |  |  |  |  |
| Improved | 1 |  | NA |  |
| Unimproved | 1.01 (0.93-1.09) | 0.804 |  |  |
| Open | 1.10 (0.96-1.26) | 0.172 |  |  |
| Others | 1.09 (0.85-1.39) | 0.488 |  |  |

\*1: Reference category, NA: Not Applicable or model not significant.

